# Effective connectivity of LSD-induced ego dissolution

**DOI:** 10.1101/2021.12.28.21268391

**Authors:** Devon Stoliker, Leonardo Novelli, Franz X. Vollenweider, Gary F. Egan, Katrin H. Preller, Adeel Razi

**Author notes:** Corresponding author Name: Devon Stoliker, Address: Monash Biomedical Imaging, 762-772 Blackburn Rd, Clayton VIC 3168. Australia. Joint Senior Authors (Katrin H. Preller, Adeel Razi).

## Abstract

Classic psychedelic-induced ego dissolution involves a shift in the sense of self and blurring of boundary between the self and the world. A similar phenomenon is identified in psychopathology and is associated to the balance of anticorrelated activity between the default mode network (DMN) – which directs attention inwards – and the salience network (SN) – which recruits the dorsal attention network (DAN) to direct attention outward. To test whether change in anticorrelated networks underlie the peak effects of LSD, we applied dynamic causal modeling to infer effective connectivity of resting state functional MRI scans from a study of 25 healthy adults who were administered 100mg of LSD, or placebo. We found that change in inhibitory effective connectivity from the SN to DMN became excitatory, and inhibitory effective connectivity from DMN to DAN decreased under the peak effect of LSD. These changes in connectivity reflect diminution of the anticorrelation between resting state networks that may be a key neural mechanism of LSD-induced ego dissolution. Our findings suggest the hierarchically organised balance of resting state networks is a central feature in the construct of self.

**Significance:** The findings can inform the parallel between the maintenance of subject-object boundary and changes to anticorrelated canonical resting state brain networks. Effective connectivity informs the hierarchical organisation of brain networks underlying modes of perception. Moreover, the anticorrelation of brain networks is an important measure of mental function. Understanding the neural mechanisms of anticorrelation change under psychedelics help identify its relationship to psychosis and its association to psychedelic assisted therapeutic outcomes.

## Introduction

Classic psychedelics are powerful substances with low toxicity that can temporarily alter brain activity and produce profound changes to consciousness (Carhart-Harris, 2018; Johnson, Richards, & Griffiths, 2008; Preller & Vollenweider, 2018; F. X. Vollenweider, 1998; F. X. Vollenweider et al., 1997). Historical use of their mind altering effects that are undergoing translation into modern clinical therapies may constitute a crucial component of their therapeutic efficacy (Roseman, Nutt, & Carhart-Harris, 2018; Yaden & Griffiths, 2020). The subjective effects of classic psychedelics are characterised by *ego dissolution* (Preller & Vollenweider, 2018; Stoliker, Egan, Friston, & Razi, 2021), described as the shift in the sense of self and a loss of boundary between the subjective and the objective world (Dittrich, 1998; Studerus, Gamma, & Vollenweider, 2010; Vollenweider and Smallridge, 2021; Grof, 1980; Lebedev et al., 2015). Ego dissolution is a validated construct (Dittrich, 1998; Matthew M. Nour, Evans, Nutt, & Carhart-Harris, 2016; Studerus et al., 2010) and is thought to involve changes to resting state network (RSN) activity (Müller, Dolder, Schmidt, Liechti, & Borgwardt, 2018; Preller et al., 2018; Stoliker et al., 2021).

Functional magnetic resonance imaging (fMRI) investigations indicate activity across the brain is functionally integrated and forms multiple RSNs (Raichle, 2015). RSNs are associated to mental activity and the balance of connectivity between them is associated to the direction of conscious attention (Michael D. Fox et al., 2005; K. J. Friston, 2011; Greicius & Menon, 2004). The default mode network (DMN) composed of the medial prefrontal cortex (mPFC), posterior cingulate cortex (PCC), and (bilateral) angular gyrus (AG) is a RSN that activates primarily in the absence of immediate external goal-directed attention (Raichle et al., 2001). Its function in self-focused thinking and attention suggests its close relationship to the ego (Andrews-Hanna, Smallwood, & Spreng, 2014). In contrast, the dorsal attention network (DAN) is a RSN activated during external-focused task-driven attention (Corbetta & Shulman, 2002; Michael D. Fox & Raichle, 2007) and is usually considered to be composed of the frontal eye field (FEF) and intraparietal sulcus (IPS) bilaterally. The activity of the DMN and DAN are identified as anticorrelated and predictably alternate with the inward or outward switching of attention (Michael D. Fox et al., 2005). The DMN-DAN anticorrelation can be hypothesised to be a mechanism maintaining the boundary between the subject (observer) and object (observation) that is altered during experiences of psychedelic ego dissolution.

A third resting state network, the salience network (SN), acts as the switching mechanism coordinating the direction of attention between internal and external stimuli (Liang et al., 2015; Seeley et al., 2007). The SN’s cardinal regions are the dorsal anterior cingulate cortex (dACC) and anterior insula (AI). Both the dACC and AI are consistently coactivated across cognitive tasks (Swick, Ashley, & Turken, 2011), however the dACC is more involved in response selection and conflict monitoring (Ide, Shenoy, Yu, & Li, 2013; Menon, 2011) while the AI receives greater multimodal sensory input (Averbeck & Seo, 2008) (Vogt & Pandya, 1987), detects behaviourally relevant stimuli (Menon, 2015) and coordinates the dynamic interactions of anticorrelated networks (Menon & Uddin, 2010; Sridharan, Levitin, & Menon, 2008).

Coordinated interactions between these networks produce important biopsychological functions. SN coactivation with the DAN detects bottom-up features in the visual environment that are infrequent or biologically significant (Egner et al., 2008; Fecteau & Munoz, 2006; Szczepanski, Pinsk, Douglas, Kastner, & Saalmann, 2013) and also enables the detection of resources relevant to higher-order goals (Menon, 2015). Furthermore, anticorrelated function between the SN and DMN is a biomarker of efficient cognition (Chand, Wu, Hajjar, & Qiu, 2017; Putcha, Ross, Cronin-Golomb, Janes, & Stern, 2016). Trauma to the white matter tracts within the SN that connect the rAI and dACC predicts dysregulated DMN function (Bonnelle et al., 2012). Importantly, abnormality of SN connectivity and its anticorrelated interactions is indicative of schizophrenia (Manoliu et al., 2014), psychosis (Palaniyappan & Liddle, 2012; Wotruba et al., 2013) and internalizing disorders (Peterson, Thome, Frewen, & Lanius, 2014) (See Menon 2015 for review of SN associations to psychopathology) (Menon, 2011).

The DMN and SN have been previously investigated in relation to unique senses of self. The SN has been suggested to be involved in an aspect of the self defined as the basic sense of being rooted within a body, termed the *minimal* or *embodied* self (Blanke & Metzinger, 2009; Lebedev et al., 2015; Legrand & Ruby, 2009). This association is supported by changes to the SN documented in psychopathology (Liu et al., 2018), meditation (Doll, Hölzel, Boucard, Wohlschläger, & Sorg, 2015; Ramirez Barrantes et al., 2019) and psychedelic-induced ego dissolution (Lebedev et al., 2015) (see Supplementary Table 1 for a subset of psychedelic findings related to networks and regions of interest). The pre-reflective qualities that define the minimal self have also been suggested as antecedents of the *narrative* aspect of self (Ho, Preller, & Lenggenhager, 2020; M. M. Nour & Carhart-Harris, 2017). The narrative aspect of self is believed to be under the control of the DMN and describes self-related mental activity and personal identity (Metzinger, 2003; Millière, 2017) that strongly parallels the classic Freudian construct of ego (Carhart-Harris & Friston, 2010; Cieri & Esposito, 2019). These parallels have led to exploratory investigations of the DMN under psychedelic-induced ego dissolution that indicate a general pattern of reduced connectivity (Carhart-Harris & Friston, 2010; Cieri & Esposito, 2019; Ruban & Kolodziej, 2018; Franz X. Vollenweider & Preller, 2020).

Reduced DMN activity has also been identified in meditation and is a feature of improved mental health (Batchelor, 2008; Judson A. Brewer et al., 2011; Hasenkamp, Wilson-Mendenhall, Duncan, & Barsalou, 2012; Millière, Carhart-Harris, Roseman, Trautwein, & Berkovich-Ohana, 2018). Psychedelics have been reported to reduce symptoms of patients experiencing internalising mental health disorders (Carhart-Harris et al., 2018; Nutt & Carhart-Harris, 2020; Ruban & Kolodziej, 2018). Altered self-boundaries may be important to these therapeutic outcomes (Griffiths et al., 2016; Roseman et al., 2018; Ross et al., 2016). Nascent alignment between the free energy principle and psychoanalytic processes proposes that high-order, secondary processes of the ego function to minimise free energy of lower-orders (Cieri & Esposito, 2019; Karl Friston, 2009; K. Friston, Kilner, & Harrison, 2006; Herzog et al., 2020). Psychedelic-induced free energy increase in networks such as the DMN is suggested to disinhibit high order inhibitory selection of bottom-up channels and enable consideration of alternate hypotheses underlying making sense of the world (Carhart-Harris & Friston, 2019; Safron, 2021; Stoliker et al., 2021). These mechanisms may broadly account for experiences of ego dissolution (Stoliker, Egan, & Razi, 2021). An early interpretation of psychedelics noted in *filtration theory* similarly suggests improvements follow psychedelic reduction of ego-protective defenses (Connolly, 2018; Freud, 1894; Stoliker et al., 2021; Swanson, 2018). Understanding network changes in ego dissolution may inform how network interactions relate to clinical outcomes. Meditation is also recognised for its benefits to wellbeing and resembles a similar trajectory of ego disarmament by seeking cessation of the self (Batchelor, 2008; Millière et al., 2018). The practice of meditation has demonstrated clinical utility (Goyal et al., 2014) and can enhance wellbeing by altering the relationship between self and other (Dambrun, 2017). Pertinently, a form of meditation termed *nondual awareness* meditation reduces the anticorrelation of extrinsic and intrinsic activated brain regions and produces a subjective experience of dissolving subject-object boundaries (Josipovic, Dinstein, Weber, & Heeger, 2012). These findings indicate anticorrelation under control of the SN as a neural mechanism that underlies ego dissolution.

Anticorrelation between the DMN and task positive networks under psilocybin has previously been investigated with the findings demonstrating reduced anticorrelation when participants experienced ego dissolution, under psilocybin (intravenous infusion, 2mg dissolved in 10ml) (Carhart-Harris et al., 2012). However, a similar investigation, under ayahuasca (oral brew, 2.2. mL/kg body weight, containing 0.8 mg/ml DMT and 0.21 mg/ml harmine) failed to identify anticorrelation changes (Palhano-Fontes et al., 2015). The inability of functional connectivity analyses to determine the direction of connectivity between networks in these studies suggests the value of adopting mechanistic approaches to determine changes in effective connectivity of networks under psychedelics. Dynamic Causal Modeling (DCM) is a Bayesian method of inference based on task based or resting state fMRI time series activity of brain regions (K. J. Friston, Harrison, & Penny, 2003; Karl J. Friston, Kahan, Biswal, & Razi, 2014). DCM can disentangle hierarchical RSN and regional interactions by determination of the directionality of connectivity. DCM has been previously applied to investigate thalamic connectivity to the cortex under LSD (Preller et al., 2019) that indicated the SN is at the apex of the DMN and DAN triple network hierarchy (Zhou et al., 2018). The SN’s position in this hierarchy and its mediating role to control the switching of DAN and DMN activity suggests that change to the SN by psychedelics may influence their patterns of anticorrelated activity. The SN and DMN share associations to aspects of self and the importance of their connectivity in psychopathology suggests change in their connectivity may be a mechanism of ego dissolution that underlies a shift in the sense of self (Northoff et al., 2006; Scalabrini et al., 2020).

Therefore, to understand the neural mechanisms of ego dissolution and inform the biological basis of the subject-object relationship, the directed changes to these networks under the classic psychedelic lysergic acid diethylamide (LSD) was investigated. LSD effects were examined across placebo (two weeks apart from LSD administration), peak effects at 75 minutes and later effects at 300 minutes post LSD administration using DCM analysis to reveal regional and network connectivity changes. Ego dissolution (quantified as oceanic boundlessness (OBN)) was measured on the five dimensions of altered states of consciousness scale (5D-ASC) (Studerus et al., 2010). Based on previous research that associated the networks under investigation to self and subject-object boundaries, we hypothesised the association between effective connectivity would change during the peak effects of LSD and ego dissolution. DMN-DAN was tested for change to subject-object boundaries and SN-DMN was tested for change to the self. However, the direction of excitatory-inhibitory connectivity change remained exploratory. Additionally, we measured connectivity of regions composing the networks of interest and the (hierarchical) connectivity strength between networks.

## Results

### Acute Drug Effects

#### Functional connectivity changes

Functional connectivity across groups demonstrated a fading of the pattern of anticorrelation between DMN and DAN and SN, from placebo to peak effects of LSD before showing evidence of restoration in the later effects at 300 minutes. Group level regional functional connectivity matrix across each condition (placebo, 75 minutes after LSD administration and 300 minutes post LSD administration) is provided in Supplementary Fig. 1.

#### Between networks changes in effective connectivity

The first resting-state fMRI scan was done 75 minutes post administration of LSD which is during the peak effects of LSD. The connectivity strength between networks was computed as the difference between averaged and unsigned efferent and afferent connection parameters between networks (see methods for further details) (Zhou et al., 2018). As shown in Fig. 1a), at this time, group level, between networks, effective connectivity increased from the SN to DMN causing the directed connection to become excitatory. A similar change was observed in the excitatory connectivity from the DMN to the DAN resulting in reduced inhibitory connectivity (see Fig. 1c). SN to DMN and DMN to DAN changes from placebo are greatest during the peak effects at 75 minutes and reduce in the later effects at 300 minutes (see Fig. 1a) and 1b). These changes show reduced hierarchical connectivity strength (i.e., increased afferent connections) of the SN and increased hierarchical connectivity strength (i.e., increased efferent connections) of the DMN and DAN from placebo (SN= −1.16 Hz, DMN = + 0.32 Hz, DAN = +0.41 Hz, see Supplementary S4 for explanation and quantitative analysis).

**Fig 1.**
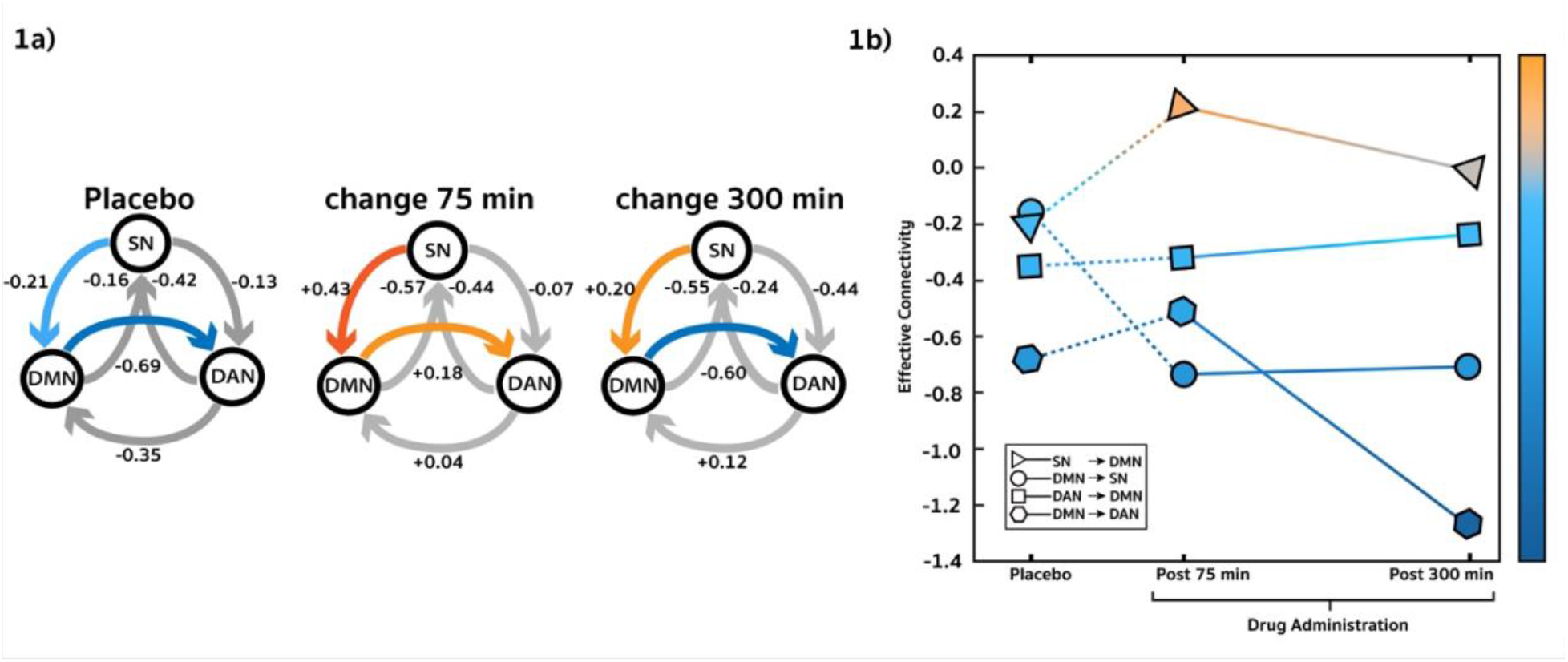
Network effective connectivity change under peak effects of LSD. a) Highlighted connections show changes in effective connectivity compared to placebo signifying peak effect. b) Network effective connectivity change graphed across placebo, peak effects and later effects. Same data as a) and b) but plotted as a line graph for better visualization. Values display effect sizes (posterior expectations) of connections in Hz. All results are for posterior probability > 0.99.

#### Between regions changes in effective connectivity

Between regions effective connectivity that underly the SN to DMN changes include increased inhibitory self-connectivity of the dACC and lFEF and effective connectivity from the lAG to rAG, lFEF to the lIPS, lIPS to rAI. These regions are highlighted because the change in their effective connectivity show large effect sizes during the peak effects than in later effects of LSD. Moreover, lAI to dACC and rAG to rAI effective connectivity show large effect size change from inhibitory in the peak effects of LSD to excitatory in the later effects (see Fig. 2, panel a) below and Supplementary Fig. 3 for untranslated effective connectivity matrices across conditions).

**Fig 2.**
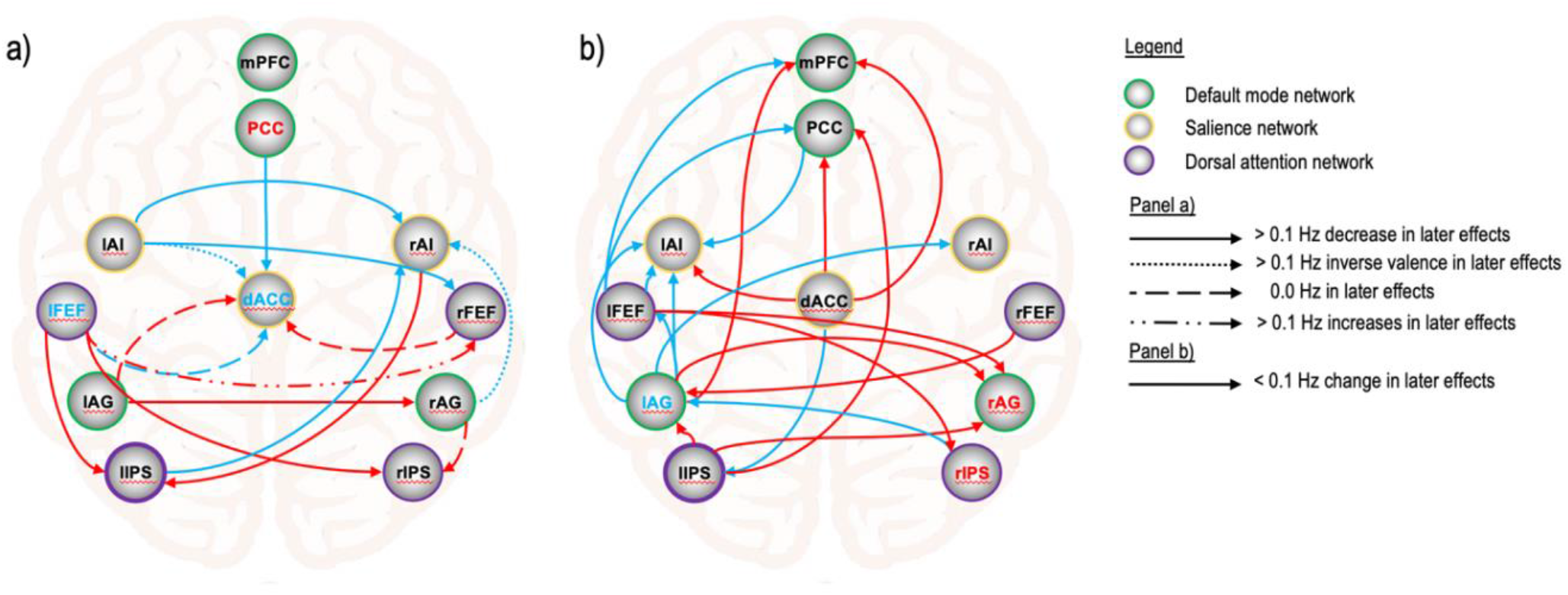
Region effective connectivity signifying a) peak and b) lasting effects of LSD. Red lines and lettering indicate excitatory change; blue lines and lettering indicate inhibitory change. Only connections with change from placebo to 75 minutes post LSD > 0.1 Hz are included. Panel a) illustrates effective connectivity with change > 0.1 Hz from peak effects at 75 minutes to the later effects at 300 minutes, signifying connectivity unique to the peak effects of LSD. See Supplementary Table 2 for posterior expectations (effect sizes) and credible intervals. Panel b) illustrates effective connectivity with change < 0.1 Hz from peak effects at 75 minutes to the later effects at 300 minutes, signifying connectivity changes that last over the effects of LSD. See Supplementary Table 3 for posterior expectations (effect size) and credible intervals. All results are for posterior probability > 0.99.

### Lasting effects

#### Between networks changes in effective connectivity

LSD effects are also distinguished by changes from placebo that last across time under LSD. Increased DAN to DMN and decreased DMN to SN effective connectivity at 75 minutes remains evident 300 minutes after LSD (see Fig. 1, panel a) above, and see Supplementary Fig. 3 for effect size and posterior probabilities).

#### Between regions changes in effective connectivity

Effects lasting across time under LSD are identified by change in lAG and rIPS self-inhibition, lFEF to mPFC inhibition and lIPS to PCC excitation. These connections show large effect sizes from placebo to peak effect that remain to the later effects of LSD (see Fig. 2, panel b) above and Supplementary Fig. 3 for untranslated effective connectivity matrices across conditions).

The results demonstrate increased effective connectivity of the DMN to DAN and SN to the DMN during the peak effects of LSD. These changes correspond with reduced SN hierarchical connectivity strength (i.e., increased afferent connections) and coincide with a fading of the functional anticorrelation (see Supplementary Fig. 1 for functional connectivity results).

## Discussion

This investigation seeks to understand how effective connectivity between anticorrelated large-scale brain networks is related to ego dissolution. Our analysis reveals between network effective connectivity changes that occur with a diminution of the pattern of anticorrelation under the peak effects of LSD. Bidirectional changes in effective connectivity between the DMN and DAN were investigated for their relationship to subject-object boundaries. We identified reduced inhibition of the DMN to the DAN under the peak effects of LSD. The reduced inhibition is largely lost in the later effects, when ego dissolution dissipates. This indicates reduced inhibition of the DMN to DAN as a feature of peak LSD effects that may relate to the fading of the functional anticorrelation between the two networks. Reduced DMN to DAN inhibition may also represent an increased transmission and connection of the narrative self to the sense of object. Moreover, hierarchical organisation and strength of networks were calculated using efferent vs afferent connections (Zhou et al., 2018). The DMN and DAN showed increased hierarchical connectivity strength during the peak effects of LSD and segregate from SN, under the peak effects of LSD (See Supplementary S4). The increase in hierarchical connectivity strength of the DMN and DAN reinforces evidence of their fading anticorrelation that may contribute to the dissolution of the boundary between the subject and the object. The opposite effective connectivity from DAN to DMN also displays reduced inhibition under LSD. However, this change remains over the course of time suggesting it is not a primary mechanism of the reduced functional anticorrelation between them or ego dissolution.

Network level DMN-DAN changes are supported by the lAG to rAG connectivity. The AG serves functions in autobiographical memory and bodily awareness (Bréchet, Grivaz, Gauthier, & Blanke, 2018). AG also operates complex language functions and makes meaning out of visually perceived self-related words (Yaoi, Osaka, & Osaka, 2015). Excitatory change in the connection from lAG to rAG in placebo to the peak effects of LSD is greater than in the later effects suggesting the importance of this connection in the peak effects.

Inclusion of the SN in this analysis enabled measurement of its effective connectivity to the DMN and DAN under LSD. The change to the coordinated balance of networks, under the control of SN, by LSD may be an important but overlooked neural mechanism of ego dissolution suggested by the superiority of SN in this hierarchy of triple networks (Zhou et al., 2018). Previous reports of reduced anticorrelation (Carhart-Harris et al., 2012), reduced SN integrity (Lebedev et al., 2015) in occurrences of ego dissolution, and its hypothesised function in basic conscious-awareness also indicate the value of measuring SN connectivity in the anticorrelation between DMN and DAN. Moreover, ego dissolution has previously been suggested to involve the breakdown of sub-personal processes underlying the minimal self, a suggestion that is consistent with Bayesian models of phenomenal selfhood in which the subjective structure of conscious experience is determined from the optimisation of prediction in perception and action (Clark, 2013; Karl Friston, 2010; Millière, 2017). The change in the SN effective connectivity under the peak effects of LSD is our most pronounced finding. SN connectivity to the DMN changes from inhibitory to excitatory before returning to inhibitory in the later effects. This flip of valence suggests SN connectivity change to the DMN is mechanistic in the peak effects of LSD and may indirectly influence the DMN-DAN interactions. The opposite connection, the DMN to SN, shows an inverse pattern of change from placebo, suggesting reduced DMN influence over the SN which lasts over time. The SN to DMN connectivity change may therefore be a more likely mechanism of ego dissolution representing a quietening of narrative self in the peak effects of LSD that reduces in the later effects. This change in DMN function to minimise free energy and suppress prediction errors that in Freudian terms may liberate the ego from the reality principle (Carhart-Harris & Friston, 2010; Cieri & Esposito, 2019). SN to DMN change is accompanied by reduced hierarchical connectivity strength of the SN that is characterised by increased SN afferent connections and increased DMN efferent connections. The SN has previously been identified to be hierarchically superior to the DMN and DAN (Zhou et al., 2018). We demonstrate the divergence of SN and DMN hierarchical strength under the peak effects of LSD shift the hierarchical order of the SN beneath the DMN.

The changes to region level effective connectivity underlying the SN are demonstrated by dACC self-connectivity under the peak effects of LSD. The dACC is a central brain hub involved in cognitive functions, social emotions (Andrews-Hanna et al., 2014; Etkin, Egner, & Kalisch, 2011; Preller et al., 2015) and performance monitoring (Ham et al., 2014) and has previously been noted in social cognition under psilocybin (Preller et al., 2016). Enlargement of the ACC has also been reported after regular use of ayahuasca (Carlos Bouso et al., 2015). We find dACC self-connectivity inhibition and inhibitory connectivity from the lAI to the dACC under the peak effects of LSD, compared to placebo. In the later effects dACC inhibition become less inhibitory while the lAI changes to excitatory. The AI is involved in interoceptive awareness (Chong, Ng, Lee, & Zhou, 2017) and the shift of internal and external perception (Menon & Uddin, 2010; Sridharan et al., 2008). Damage to SN connectivity predicts dysregulated DMN function (Menon, 2011) and alteration of SN anticorrelated interactions share association with psychosis and internalising disorders (Bonnelle et al., 2012). dACC and lAI inhibition at the peak effects of LSD may indicate the role of SN inhibition that is relevant to ego dissolution and psychedelic assisted therapy. Connectivity between the DMN and SN is also noted in region changes from the rAG to rAI. This connection follows the lAI to dACC pattern of inhibition in the peak effects and excitation in the later effects, suggesting a reduced influence of the DMN to the SN during peak effects, when ego dissolution occurs that may suggest a shifting sense of self.

The PCC and dACC represent cardinal regions of the DMN and SN respectively and are related to the narrative and minimal aspects of self (Lebedev et al., 2015; Millière, 2017). The connectivity changes between them are reported in association to meditation and the mindfulness capacities of meditators (Judson A. Brewer et al., 2011; Millière et al., 2018; Palhano-Fontes et al., 2015). We provided evidence of the excitation from the dACC to PCC in our effective connectivity analysis under peak effects (see Supplementary Fig. 3), which may suggest dACC cognitive control functions influence on self-related functions of PCC. Interestingly, self-connectivity of the PCC, a hub region of DMN, increases excitation during the peak effects of LSD. PCC involvement in self-awareness is well documented (J. A. Brewer, Garrison, & Whitfield-Gabrieli, 2013; Northoff et al., 2006) and activity of this region is commonly observed to decrease under psychedelics (see Supplementary Table 1).

Other notable regions in our analysis include the FEF and IPS. The FEF operates visual attention (Thompson, Biscoe, & Sato, 2005) and the IPS serves functions directing attention and memory (Corbetta & Shulman, 2002; Michael D. Fox, Corbetta, Snyder, Vincent, & Raichle, 2006). The nearby right inferior parietal lobule (rIPL) is also associated with the sense of agency and self-other discrimination (Chaminade & Decety, 2002; Lucina Q. Uddin, Molnar-Szakacs, Zaidel, & Iacoboni, 2006). Smaller effect sizes involving these regions were identified in our analysis (see Supplementary Fig. 3) suggesting more subtle widespread brain changes occur under the effects of LSD.

Taken together, under the peak effects of LSD, a strong increase in change of effective connectivity, and a widening of gap between the hierarchical connectivity strength of the SN vs the DMN and DAN shifts in a direction antithetical to normal hierarchical organisation. This may be said to resemble a collapse – or flattening – of the hierarchy, during peak effects of LSD, when ego dissolution occurs (See Fig. 1 and Supplementary S4). Effective connectivity explains this effect as increased excitatory connectivity from the SN to the DMN. These directed connection changes and the hierarchical strength changes may function to alter the relationship between the minimal and narrative senses of self. The direction of this change suggests the influence of the minimal-self traverses over the narrative-self and may relate to the shift in sense of self described under ego dissolution.

Modelling the connectivity of ego dissolution can provide a means to determine the neural mechanisms that underlie the perception of inner and outer reality. The present research establishes important steps to identify the change in network interactions associated with the dichotomy of the subject-object relationship under LSD. The networks involved in this interaction are important in cognitive function and mental wellbeing, suggesting that understanding their change due to psychedelics may elucidate mechanisms of psychedelic clinical therapy. Consideration of practical and theoretical challenges may aid future research directed to study ego dissolution and the anticorrelation between brain networks.

Our relatively small sample size (n=20) is clearly a limitation. Sample size and processing pipeline strongly impact the reliability of results. Small sample size may also account for unexpected placebo effective connectivity in our group of subjects. A second practical limitation is the large variance of participant subjective responses to a standard dose of LSD (100mg). Averaging the connectivity of participants experiencing highly variable subjective shifts in consciousness may dilute the effective connectivity representing LSD’s subjective effects. An alternative to increased sample size may be predetermining participant dose-response and including only participants with high subjective responses in the analysis. Replicating the investigation of anticorrelated networks with alternate classic psychedelic substances including psilocybin, ayahuasca and mescaline in addition to LSD will be a worthy direction to validate our results. Furthermore, comparisons with similar altered forms of consciousness reporting ego change such as psychosis and meditation may also help discern the function of components in the anticorrelated networks.

The selection of regions and region coordinates composing the networks of interest require careful attention. For example, some studies define the DAN as composed by the posterior prefrontal cortex, the inferior precentral sulcus, the superior occipital gyrus, the middle temporal motion complex, and the superior parietal lobule (Andrews-Hanna et al., 2014). Similarly, the DMN may be composed using the inferior parietal lobule (Di & Biswal, 2014) closely situated to the IPS used in the DAN in this analysis. The composition of regions composing networks can strongly impact results. Furthermore, selection of region centroids can also affect the results. Previous work using this LSD data set used different coordinates for the PCC and identified its increased self-inhibition (Preller et al., 2019), although our analysis identified decreased PCC inhibition. Avoiding this problem is challenging due to anatomical variability between participants, variability in the method of determining coordinates and the selection of regions composing networks. However, the standardisation of these considerations across studies is important to improve the accuracy and reliability of findings across studies.

Future work to extend the current scope of analysis to include connectivity dynamics of additional task positive networks anticorrelated to the DMN and under control of the SN is required. For example, non-psychedelic research involving the CEN (central executive network, also known as the frontoparietal central executive network (FPCEN)) has been conducted to investigate schizophrenia (Leptourgos et al., 2020; Menon, 2011), meditation (Doll et al., 2015) and control of attention (Corbetta & Shulman, 2002). Its importance is further signified by investigations of large-scale network interactions and anticorrelations with the DMN (Andrews-Hanna et al., 2014; Bolton et al., 2020; Menon, 2018). Inclusion of the CEN in anticorrelation investigations may provide a more complete account of anticorrelated network changes associated to ego dissolution.

Theoretical challenges include consensus on the nature and situation of ego dissolution within the taxonomy of psychedelic-induced effects. Psychometric investigations of psychoanalytic based concepts of ego-functions and measurement of their brain connectivity are recommended future directions. Psychedelics may contribute to these investigations by altering and allowing measurement of brain mechanisms hypothesised to underlie psychoanalytic constructs. Integration of psychoanalytic theory and brain connectivity processes may be important to identifying processes involved in enabling the sense of self and experiences of ego dissolution.

## Conclusions

The between network balance of anticorrelated activity of specific RSNs depends on subtle adjustments to network activation. Mental wellbeing and efficient cognition rely on the balance of these interactions. LSD appears to shift the balance of network activation and diminish the anticorrelation between brain networks responsible for internal and external modes of perception. Observed increases in effective connectivity from the DMN to DAN and increased hierarchal strength of these networks under peak effects may account for the blurring of boundary between subject and object experienced in ego dissolution. Ego dissolution also involves a shift in the sense of self that may be explained by changes to the interactions between SN and DMN. These networks are related to distinct aspects of self. Increased effective connectivity from the SN to the DMN and decreased SN hierarchical connectivity strength emphasise hierarchical flattening and increased salience functions reaching the DMN under the peak effects of LSD. Future research could model how changes to these networks effective connectivity may support psychedelic therapeutic outcomes. The neuroscientific study of psychedelic-induced ego dissolution reminds us that constructs and representations of self and internal and external reality exist in connectivity dynamics. This intriguing understanding may inspire future investigations of sentience and consciousness to learn how normal brain function mechanisms contribute to the subject-object relationship and frame our perspective of reality.

## Methods

### Participants

The data analysed in this paper were collected as part of a larger study (registered at ClinicalTrials.gov (NCT02451072)), which is reported in (Preller et al., 2017) and (Preller et al., 2018), and was approved by the Cantonal Ethics Committee of Zurich. 25 subjects (19 males and 6 females; mean age = 25.24 y; SD = 3.72 y; range = 20–34 y) were recruited through advertisements at universities in Zurich, Switzerland. All participants were deemed healthy after screening for medical history, physical examination, blood analysis, and electrocardiography. The Mini-International Neuropsychiatric Interview (MINI-SCID), the Diagnostic and Statistical Manual of Mental Disorders, fourth edition self-rating questionnaire for Axis-II personality disorders (SCID-II), and the Hopkins Symptom Checklist (SCL-90-R) were used to exclude subjects with present or previous psychiatric disorders or a history of major psychiatric disorders in first-degree relatives. Participants were asked to abstain from prescription and illicit drug use two weeks prior to first testing and throughout the duration of the study, and abstain from alcohol use 24 hours prior to testing days. Urine tests were also used to exclude pregnancy. Further exclusion criteria included left-handedness, poor knowledge of the German language, cardiovascular disease, history of head injury or neurological disorder, history of alcohol or illicit drug dependence, MRI exclusion criteria, including claustrophobia, and previous significant adverse reactions to a hallucinogenic drug. All participants provided written informed consent statements in accordance with the declaration of Helsinki before participation in the study. Subjects received written and oral descriptions of the study procedures, as well as details regarding the effects and possible risks of drug treatment.

### Design

A double blind, randomized, placebo-controlled, cross-over study was performed. Testing days occurred two weeks apart and participants were orally administered either LSD after pretreatment with 179 mg Mannitol and 1 mg Aerosil (LSD condition) or 179 mg Mannitol and 1 mg Aerosil after pre-treatment with 179 mg Mannitol and 1 mg Aerosil (placebo condition). Resting state scans (10 minutes each) were taken 75 and 300 minutes following administration.

### MRI Data Acquisition and Preprocessing

MRI data were acquired on a Philips Achieva 3.0T whole-body scanner. A 32-channel receive head coil and MultiTransmit parallel radio frequency transmission was used. Images were acquired using a whole-brain gradient-echo planar imaging (EPI) sequence (repetition time, 2,500 ms; echo time, 27 ms; slice thickness, 3 mm; 45 axial slices; no slice gap; field of view, 240 × 240 mm^2^; in-plane resolution, 3 × 3 mm; sensitivity-encoding reduction factor, 2.0). Additionally, high-resolution anatomical images (voxel size, 0.7 × 0.7 × 0.7 mm^3^) were acquired using a standard T1-weighted 3D magnetization prepared rapid-acquisition gradient echo sequence. The acquired images were analysed using SPM12 (https://www.fil.ion.ucl.ac.uk). The pre-processing steps of the images consisted of slice-timing correction, realignment, spatial normalization to the standard EPI template of the Montreal Neurological Institute (MNI), and spatial smoothing using a Gaussian kernel of 6-mm full-width at half maximum. Head motion was investigated for any excessive movement, but movement did not exceed 3 mm in any participant.

### Extraction of region coordinates across subjects

Group ICA for fMRI Toolbox (GIFT, http://mialab.mrn.org/software/gift) (Calhoun et al. 2001) was used to identify the three resting state networks of interest from placebo scans. Pre-processed resting state fMRI data were spatially sorted into 20 components (Biswal et al. 2010; Shirer et al. 2012; Tsvetanov et al. 2016) and spatially matched with pre-existing network templates (Shirer et al. 2012).

Networks were composed of cardinal regions constituting a core part of DMN (Andrews-Hanna, Reidler, Sepulcre, Poulin, & Buckner, 2010; Dixon et al., 2017) which reliably show anticorrelation with the DAN and SN (Chen, Glover, Greicius, & Chang, 2017; Dixon et al., 2017; M. D. Fox, Zhang, Snyder, & Raichle, 2009; Fransson, 2005; L. Q. Uddin, Kelly, Biswal, Castellanos, & Milham, 2009) and followed the selection of regions in a related investigation by Zhou and colleagues (Zhou et al., 2018).

Identification of cardinal nodes within each intrinsic network – averaged across our subjects – was located using peak RSN activity of clusters within networks (p=.05) visualized using xjView toolbox (https://www.alivelearn.net/xjview). Associations between peak coordinates and cardinal nodes of network regions of interest (ROI) were determined by expert visual inspection. The MNI coordinates of the selected ROIs are listed in Table 4.

**Table 4.** Coordinates of regions of interest. The DMN comprised of the posterior cingulate cortex (PCC), medial prefrontal cortex (mPFC), and left and right angular gyrus (lAG/rAG); the SN comprised of the dorsal anterior cingulate cortex (dACC), left and right anterior insula (lAI/rAI); and the DAN comprised the left and right frontal eye field (lFEF/rFEF) and the left and right inferior parietal sulcus/superior parietal lobule (lIPS(SPL)/rIPS(SPL).

| <u>Region</u> | <u>MNI coordinates</u> |  |  | <u>Network</u> |
| --- | --- | --- | --- | --- |
|  | x | y | z |  |
| PCC | 0 | -73 | 38 | DMN |
| mPFC | 0 | 53 | 32 | DMN |
| lAG | -54 | -61 | 29 | DMN |
| rAG | 57 | -58 | 20 | DMN |
| dACC | 0 | 17 | 41 | SN |
| lAI | -48 | 14 | -7 | SN |
| rAI | 48 | 14 | -7 | SN |
| lFEF | -30 | -10 | 68 | DAN |
| rFEF | 30 | -7 | 68 | DAN |
| lIPS/SPL | -42 | -43 | 62 | DAN |
| rIPS/SPL | 39 | -40 | 65 | DAN |

A generalized linear model (GLM) was used to regress 6 head motion parameters (3 translation and 3 rotational), white matter and cerebrospinal fluid signals from preprocessed data. One subject was excluded as no activation was found in one or more regions of interest. We also used global signal regression in our pre-processing pipeline. The time series for each ROI was computed as the first principal component of the voxel activity within a 6 mm sphere centred on the ROI coordinates (as listed in Table 4).

### Anticorrelation Functional Connectivity Validation

The functional connectivity matrix between regions of the DMN, SN, and DAN in the placebo condition was computed for all subjects. Since our arguments rest on the premise of functional anticorrelation between the resting-state networks (and similar second-order features are used for model estimation by the following spectral DCM analysis), 4 subjects that did not show evidence of the anticorrelation in the placebo condition were excluded from further analysis. As such, 20 subjects were left available for the following DCM analysis.

### Specification and Inversion of DCM

A fully-connected DCM was specified using the 11 ROIs defined in Table 4, without any exogenous inputs. The DCM for each subject was then inverted using spectral DCM (Karl J. Friston et al., 2014; Razi, Kahan, Rees, & Friston, 2015) to infer the effective connectivity that best explains the observed cross-spectral density for each subject. This procedure was repeated for each of the three testing conditions. The DCM fitted the data very well and explained variance was over 85% across all subjects, and averaged 91%.

### Second Level Analysis Using Parametric Empirical Bayes

The effective connectivity inferred by spectral DCM for each subject are taken to the second (group) level to test hypotheses about between-subject effects. A General Linear Model (GLM) is employed to decompose individual differences in effective connectivity into hypothesised group-average connection strengths plus unexplained noise. Hypotheses on the group-level parameters are tested within the Parametric Empirical Bayes (PEB) framework (K. J. Friston et al., 2016), where both the expected values and the covariance of the parameters are taken into account. That is, precise parameter estimates influence the group-level result more strongly than uncertain estimates, which are down-weighted. Bayesian model reduction (BMR) is used as an efficient form of Bayesian model selection (K. J. Friston et al., 2016).

### Network Level Effective Connectivity and Hierarchical Organization

The expected network-level connectivity was computed as the sum of the expected effective connectivity values between the corresponding ROIs. Then, following Zhou et. al., 2018 (Zhou et al., 2018) the hierarchical connectivity strength of each network was obtained by computing the difference between its averaged efferent and afferent connections (i.e., absolute values, see Supplementary S4). A similar approach was used for analysing hierarchical projections in the monkey brain (Goulas, Uylings, & Stiers, 2012) and prefrontal cortex hierarchical organization in humans (Nee & D’Esposito, 2016; Zhou et al., 2018).

## Data Availability

The extracted time series for each ROI and each condition, as well as all statistics applied to these data, have been deposited in Bitbucket. See
Preller KH, et al. (2019) data from Effective connectivity changes in LSD-induced altered states of consciousness in humans, available at https://bitbucket.org/katrinpreller/effective-connectivity-changes-in-lsd-induced-altered-states. Deposited January 9, 2019.

## Data Availability

All data produced in the present work are contained in the manuscript.

