## Supplementary material for "Effective connectivity of LSD-induced ego dissolution"

### Neural mechanisms underlying LSD-induced ego dissolution

Devon Stoliker[1], Leonardo Novelli[1], Franz Vollenweider[3], Gary F. Egan[1,2], Katrin Preller[3,*], and Adeel Razi[1,2,4,5,*]
[1]Turner Institute for Brain and Mental Health, Monash University, Clayton, VIC
[2] Monash Biomedical Imaging, Monash University, Clayton, VIC
[3] Department of Psychiatry, Psychotherapy & Psychosomatics, University Hospital for Psychiatry, Zurich, Switzerland
[4] Wellcome Centre for Human Neuroimaging, UCL, London, United Kingdom
[5]CIFAR Azrieli Global Scholars Program, CIFAR, Toronto, Canada
*Joint senior authors

#### Methods:

#### 1. Spectral Dynamic Causal Modelling

Dynamic causal modelling (DCM) is Bayesian framework that infers the directed (causal) connectivity among the neuronal systems – referred to as effective connectivity. We recently proposed a new DCM for resting state fMRI – based upon a deterministic model that generates predicted cross spectra – referred to as spectral DCM. In order to model resting state activity – in the absence of external stimuli – we will have to add a stochastic component, i.e. neural fluctuations, to the classical DCM based on ordinary differential equations. Mathematically, we can express the formulation of the stochastic generative model using a set of two equations. First is the neuronal state equation, namely

$$\dot{x}(t) = f(x(t), u(t), \theta) + v(t), \tag{S1}$$

and second is the observation equation, which is a static nonlinear mapping from the hidden physiological states in (1) to the observed BOLD activity and is written as:

$$y(t) = h(x(t), \varphi) + e(t), \tag{S2}$$

where $\dot{x}$(t) is the rate of change of the neuronal states $x(t)$, $\theta$ are unknown parameters (i.e. the effective connectivity) and $v(t)$ (resp. $e(t)$) is the stochastic process – called the state noise (resp. the measurement or observation noise) – modelling the random neuronal fluctuations that drive the resting state activity. In the observation equations, $\varphi$ are the unknown parameters of the (haemodynamic) observation function and $u(t)$ represents any exogenous (or experimental) inputs that drive the hidden states – that are usually absent in resting state designs[1]. Spectral DCM furnishes a constrained inversion of the stochastic model by parameterising the neuronal fluctuations $v(t)$. Spectral DCM simplifies the generative model by replacing the original timeseries with their second-order statistics (i.e., cross spectra). This means, instead of estimating time varying hidden states, we are estimating their covariance which is time invariant. Then we simply need to estimate the covariance of the random fluctuations; where a scale free (power law) form for the state noise (resp. observation noise) is used – motivated from previous work on neuronal activity [2-4] – as follows:

$$g_v(\omega,\theta) = \alpha_v \omega^{-\beta_v}$$

$$g_e(\omega,\theta) = \alpha_e \omega^{-\beta_e} \qquad \text{(S3)}$$

Here, $\{\alpha,\beta\} \subset \theta$ are the parameters controlling the amplitudes and exponents of the spectral density of the neural fluctuations. The parameterisation of endogenous fluctuations means that the states are no longer probabilistic; hence the inversion scheme is significantly simpler, requiring estimation of only the parameters (and hyperparameters) of the model.

We used standard Bayesian model inversion to infer the parameters of the model in (1), (2) and (3), from the observed signal $y(t)$. The description of the Bayesian model inversion procedures based on variational Laplace can be found elsewhere for the interested readers [5-7].

#### 2. Parametric Empirical Bayes

Empirical Bayes refers to the Bayesian inversion or fitting of hierarchical models. In hierarchical models, constraints on the posterior density over model parameters at any given level are provided by the level above. These constraints are called empirical priors because they are informed by empirical data. We recently introduced a second-level or between-subjects model over parameters, which represents how individual (within-subject) connections derive from the subjects' group membership [8] – based on parametric empirical Bayes (PEB). This approach calls on Bayesian Model Reduction (BMR) to finesse the inversion of multiple models of a single dataset or a single (hierarchical) model of multiple datasets. BMR allows one to compute posterior densities over model parameters, under new prior densities, without explicitly inverting the model again. For example, one can invert a DCM for each subject in a group and then evaluate the posterior density over group effects, using the posterior densities over parameters from the single subject inversion. This may improve subject-specific parameter estimates, by using group-level estimates to rescue individual DCM from local optima. Mathematically, for DCM studies with $N$ subjects and $M$ parameters per DCM, we have a hierarchical model, where the responses of the $i$-th subject and the distribution of the parameters over subjects can be modeled as:

$$y_i = \Gamma_i^{(1)}(\theta^{(1)}) + \varepsilon_i^{(1)} \qquad \text{(S4)}$$

$$\theta^{(1)} = \Gamma^{(2)}\big(\theta^{(2)}\big) + \varepsilon^{(2)}$$

$$\theta^{(2)} = \eta + \varepsilon^{(3)}$$

where, $y_i$ is the BOLD time series from *i-th* subject *and* $\Gamma_i^{(1)}$ is a nonlinear mapping from the parameters of a model to the predicted response $y$ for e.g. as shown in Eq. S1 above. $\varepsilon_i^{(1)}$ is independent and identically distributed (i.i.d.) observation noise (equivalent to $e(t)$ in Eq. S2). In this hierarchical form, *empirical priors* encoding second (between-subject) level effects place constraints on subject-specific parameters. The second level would be a linear model where the random effects are parameterised in terms of their precision:

$$\Gamma^{(2)}\big(\theta^{(2)}\big) = (X \otimes W)\beta$$

where, $\beta \subset \theta$ are group means or effects encoded by a design matrix with between $X$ and within-subject $W$ parts. The between-subject part encodes differences among subjects or covariates such as age, while the within-subject part specifies mixtures of parameters that show random effects. We assume that the first column of the design matrix is a constant term, modelling group means and subsequent columns encode group differences or covariates such as age.

| *Author* | *Network* | *Substance* | *Finding* |
|---|---|---|---|
| Tagliazucchi et al., 2014 | DMN | psilocybin | Increased variability in BOLD signal spectral behaviour |
| Muller et al., 2018 | DMN | LSD | Decreased within functional connectivity |
| Palhano-Fontes et al., 2015 | DMN | ayahuasca | Decreased within functional connectivity |
| Tagliazucchi et al., 2016 | DMN, SN | LSD | Increased between network connectivity |
| Atasoy et al., 2017 | DMN, SN | LSD | Increased coupling associated to mood, arousal, and ego dissolution |
| Lebedev et al., 2015 | SN | psilocybin | Disintegration of SN associated to ego dissolution |
| Tagliazucchi et al., 2014 | DAN | psilocybin | Increased variability of BOLD signal spectral behaviour |
| Carhart-Harris et al., 2012 | DMN-TPN | psilocybin | Increased functional connectivity (DMN-TPN) |
| Palhanos-Fontes et al., 2015 | DMN-TPN | ayahuasca | No significant change reported between (DMN-TPN) |
| | *Region* | | |
| Preller et al., 2019 | PCC | LSD | Increased effective connectivity from thalamus to PCC |
| Carhart-Harris et al., 2012 | PCC | psilocybin | Decreased blood flow |
| Muthukumaraswami et al., 2013, Kometer et al., 2015 | PCC | psilocybin | Decreased neuronal oscillations using MEG; EEG |
| Smigelski et al., 2019 | mPFC-PCC | psilocybin | Decoupling of functional connectivity |
| | mPFC-AG | | Decreased activity associated to subjective effects |
| Carhart-Harris et al., 2012 | mPFC-PCC | psilocybin | Decoupling of functional connectivity |
| | mPFC/ACC | | Decreased activity associated to subjective effects |
| Tagliazucchi et al., 2014 | ACC | psilocybin | Increased amplitude of BOLD signal fluctuations (entropy) |
| Sampedro et al., 2012 | PCC-ACC | ayahuasca | Increased functional connectivity |

*Table S1. Psychedelic findings related to networks of interest (default mode network (DMN), salience network (SN) and dorsal attention network (DAN)).*

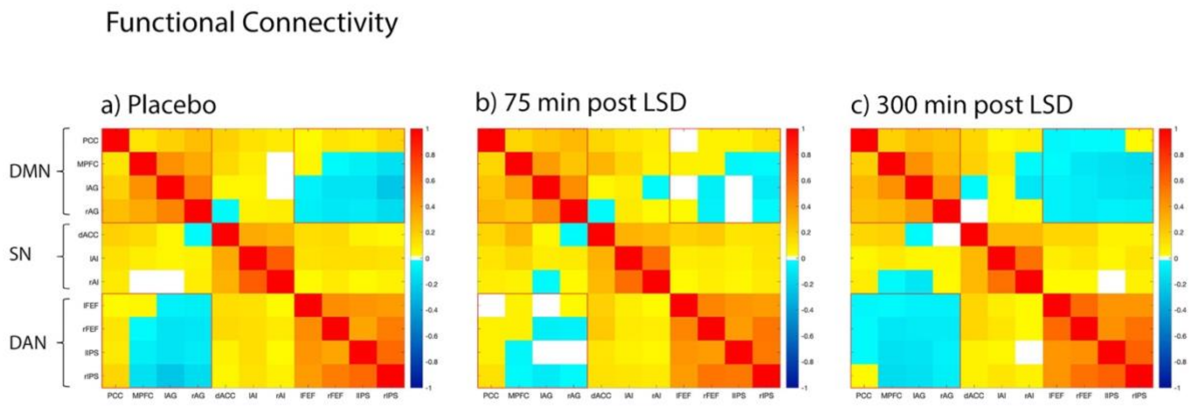


*Fig S1. Functional connectivity across conditions. Reduced anticorrelation is observed between the DMN and DAN under peak effects, 75 minutes after adminstration of LSD.*

#### 3. Design Matrix and Results

Our design matrix designated the placebo group to serve as the baseline. Regressors in our design matrix encode: 1) placebo group 2) the additive effect of being in the second group (LSD after 75 min) relative to the placebo group, and 3) the additive effect of being in the third group (LSD after 300 min) relative to the placebo group. See design matrix and results below.

$$\begin{bmatrix} 1 & 0 & 0 \\ 1 & 0 & 0 \\ 1 & 1 & 0 \\ 1 & 1 & 0 \\ 1 & 0 & 1 \\ 1 & 0 & 1 \end{bmatrix}$$

*Fig S2. Change from placebo design matrix.*

#### Region EC matrices

##### a) Placebo

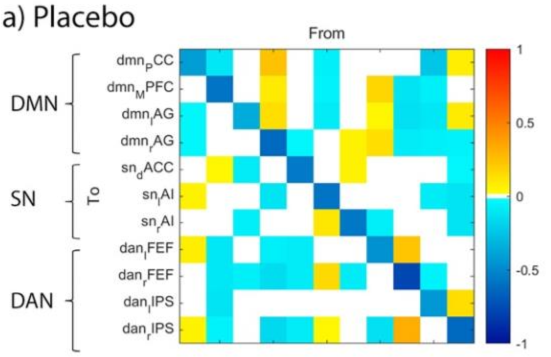


| | PCC | mPFC | lAG | rAG | ACC | lAI | rAI | lFEF | rFEF | lIPS | rIPS |
|---|---|---|---|---|---|---|---|---|---|---|---|
| PCC | -0.408 | -0.1027 | 0.000 | 0.240 | 0.000 | -0.076 | 0.000 | 0.000 | 0.000 | -0.235 | 0.073 |
| mPFC | 0.000 | -0.5827 | 0.000 | 0.087 | 0.000 | -0.053 | 0.000 | 0.162 | -0.113 | -0.077 | 0.000 |
| lAG | -0.045 | 0.000 | -0.343 | 0.139 | 0.000 | 0.083 | 0.000 | 0.043 | -0.133 | -0.102 | 0.089 |
| rAG | -0.039 | 0.000 | 0.000 | -0.634 | -0.046 | 0.000 | 0.049 | 0.132 | -0.080 | -0.064 | -0.048 |
| ACC | 0.000 | 0.043 | -0.081 | 0.000 | -0.549 | 0.000 | 0.056 | 0.000 | 0.000 | 0.000 | -0.042 |
| lAI | 0.060 | 0.000 | 0.000 | -0.109 | 0.000 | -0.591 | 0.000 | 0.000 | 0.000 | -0.083 | -0.110 |
| rAI | 0.000 | 0.000 | -0.072 | 0.000 | 0.000 | 0.104 | -0.572 | -0.072 | 0.000 | 0.000 | -0.111 |
| lFEF | 0.074 | -0.095 | 0.000 | -0.076 | -0.081 | 0.000 | 0.000 | -0.468 | 0.224 | 0.000 | 0.000 |
| rFEF | 0.000 | -0.100 | -0.066 | -0.150 | -0.079 | 0.154 | -0.086 | 0.000 | -0.776 | -0.059 | 0.000 |
| lIPS | 0.000 | -0.117 | 0.000 | 0.000 | 0.000 | 0.000 | 0.000 | 0.000 | 0.000 | -0.433 | 0.126 |
| rIPS | 0.061 | -0.058 | 0.000 | -0.162 | -0.082 | 0.046 | 0.000 | -0.138 | 0.315 | 0.000 | -0.626 |

##### b) 75 minutes

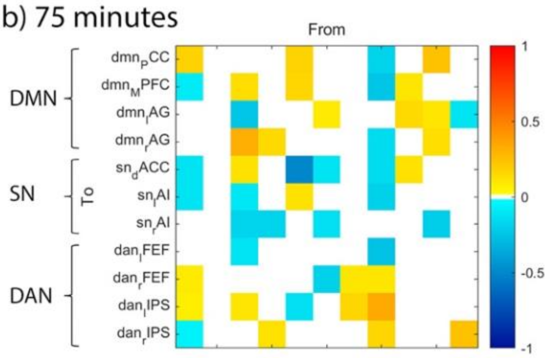


| | PCC | mPFC | lAG | rAG | ACC | lAI | rAI | lFEF | rFEF | lIPS | rIPS |
|---|---|---|---|---|---|---|---|---|---|---|---|
| PCC | 0.184 | 0.000 | 0.000 | 0.000 | 0.178 | 0.000 | 0.000 | -0.182 | 0.000 | 0.236 | 0.000 |
| mPFC | -0.079 | 0.000 | 0.138 | 0.000 | 0.161 | 0.000 | 0.000 | -0.235 | 0.095 | 0.000 | 0.000 |
| lAG | 0.000 | 0.000 | -0.249 | 0.000 | 0.000 | 0.090 | 0.000 | 0.000 | 0.151 | 0.107 | -0.116 |
| rAG | 0.000 | 0.000 | 0.319 | 0.153 | 0.000 | 0.000 | 0.000 | -0.144 | 0.000 | 0.128 | 0.000 |
| ACC | -0.116 | 0.000 | 0.116 | 0.000 | -0.509 | -0.113 | 0.000 | -0.154 | 0.112 | 0.000 | 0.000 |
| lAI | -0.110 | 0.000 | -0.109 | 0.000 | 0.123 | 0.000 | 0.000 | -0.188 | 0.000 | 0.000 | 0.000 |
| rAI | 0.000 | 0.000 | -0.177 | -0.177 | 0.000 | -0.138 | 0.000 | 0.000 | 0.000 | -0.206 | 0.000 |
| lFEF | 0.000 | 0.000 | -0.123 | 0.000 | 0.000 | 0.000 | 0.000 | -0.252 | 0.000 | 0.000 | 0.000 |
| rFEF | 0.085 | 0.000 | 0.000 | 0.000 | 0.000 | -0.199 | 0.097 | 0.106 | 0.000 | 0.000 | 0.000 |
| lIPS | 0.068 | 0.000 | 0.099 | 0.000 | -0.126 | 0.000 | 0.159 | 0.334 | 0.000 | 0.000 | 0.000 |
| rIPS | -0.061 | 0.000 | 0.000 | 0.109 | 0.000 | 0.000 | 0.000 | 0.168 | 0.000 | 0.000 | 0.237 |

##### c) 300 minutes

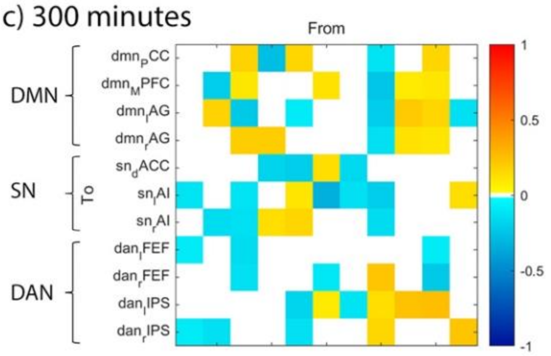


| | PCC | mPFC | lAG | rAG | ACC | lAI | rAI | lFEF | rFEF | lIPS | rIPS |
|---|---|---|---|---|---|---|---|---|---|---|---|
| PCC | 0.000 | 0.000 | 0.172 | -0.277 | 0.160 | 0.000 | 0.000 | -0.111 | 0.000 | 0.164 | 0.000 |
| mPFC | 0.000 | -0.208 | 0.105 | 0.000 | 0.000 | 0.124 | 0.000 | -0.247 | 0.086 | 0.107 | 0.000 |
| lAG | 0.000 | 0.183 | -0.228 | 0.000 | -0.089 | 0.000 | 0.000 | -0.203 | 0.209 | 0.168 | -0.135 |
| rAG | 0.000 | 0.000 | 0.195 | 0.202 | 0.000 | 0.000 | 0.000 | -0.127 | 0.119 | 0.095 | 0.000 |
| ACC | 0.000 | 0.000 | 0.000 | -0.174 | -0.204 | 0.139 | -0.168 | 0.000 | 0.000 | 0.000 | 0.000 |
| lAI | -0.110 | 0.000 | -0.115 | 0.000 | 0.096 | -0.337 | -0.126 | -0.217 | 0.000 | 0.000 | 0.130 |
| rAI | 0.000 | -0.147 | -0.137 | 0.138 | 0.161 | 0.000 | 0.000 | -0.148 | 0.000 | 0.000 | 0.000 |
| lFEF | -0.092 | 0.000 | -0.154 | 0.000 | 0.000 | 0.000 | 0.000 | 0.000 | 0.000 | -0.087 | 0.000 |
| rFEF | 0.000 | 0.000 | -0.133 | 0.000 | 0.000 | -0.097 | 0.000 | 0.227 | 0.000 | -0.233 | 0.000 |
| lIPS | 0.000 | 0.000 | 0.000 | 0.000 | -0.176 | 0.091 | -0.124 | 0.126 | 0.238 | 0.254 | 0.000 |
| rIPS | -0.089 | -0.130 | 0.000 | 0.000 | -0.137 | 0.000 | 0.000 | 0.157 | 0.000 | 0.000 | 0.221 |

*Fig S3. Group level effective connectivity (EC) across conditions a) placebo, b) 75 min and c) 300 min post LSD administration. Results in (b) and (c) represent change from placebo EC (a). Values reported in Hz. Posterior probability threshold = .99.*

*Table S2. Effective connectivity signifying the peak effects of LSD*

| | *Change from Placebo to 75 min post administration* | | *Change from Placebo to 300 min post administration* | |
|---|---|---|---|---|
| *Connection* | *Valence & Effect Size* | *Credible Intervals (low/high)* | *Valence & Effect Size* | Credible Intervals (low/high) |
| PCC → PCC | +0.184 | 0.063/0.304 | 0.000 | 0.000/0.000 |
| PCC → dACC | -0.116 | -0.153/-0.079 | 0.000 | 0.000/0.000 |
| lAG → rAG | +0.319 | 0.241/0.397 | +0.195 | 0.116/0.279 |
| lAG → dACC | +0.116 | 0.502/0.183 | 0.000 | 0.000/0.000 |
| rAG → rAI | -0.177 | -0.266/-0.087 | +0.138 | 0.053/0.222 |

| | | | | |
|---|---|---|---|---|
| rAG → rIPS | +0.109 | 0.018/0.200 | 0.000 | 0.000/0.000 |
| dACC → dACC | -0.509 | -0.672/-0.346 | -0.204 | -0.364/-0.441 |
| lAI → dACC | -0.133 | -0.165/-0.060 | +0.139 | 0.083/0.196 |
| lAI → rAI | -0.138 | -0.259/-0.175 | 0.000 | 0.000/0.000 |
| lAI → rFEF | -0.199 | -0.291/-0.108 | -0.097 | -0.191/-0.003 |
| rAI → lIPS | +0.159 | 0.099/0.219 | -0.124 | -0.186/-0.062 |
| lFEF → dACC | -0.154 | -0.222/-0.086 | 0.000 | 0.000/0.000 |
| lFEF → lFEF | -0.252 | -0.408/-0.096 | 0.000 | 0.000/0.000 |
| lFEF → rFEF | +0.106 | 0.010/0.203 | +0.227 | 0.129/0.325 |
| lFEF → lIPS | +0.334 | 0.237/0.432 | +0.126 | 0.028/0.224 |
| rFEF → dACC | +0.112 | 0.051/0.174 | 0.000 | 0.000/0.000 |
| lIPS → rAI | -0.206 | -0.297/-0.115 | 0.000 | 0.000/0.000 |

*Table S2. Changes to between regions effective connectivity signifying peak effects of LSD. Only the change from placebo > 0.1 Hz with effective connectivity change > 0.1Hz in the later effects at 300 min, compared to 75 minutes, are reported. Values reported in Hz. All results are for posterior probability > 0.99.*

*Table S3. Effective connectivity signifying the lasting effects of LSD*

| | *Change from Placebo to 75 min post administration* | | *Change from Placebo to 300 min post administration* | |
|---|---|---|---|---|
| *Connection* | *Valence & Effect Size* | *Credible Intervals (low/high)* | *Valence & Effect Size* | *Credible Intervals (low/high)* |
| PCC → lAI | -0.110 | -0.165/-0.055 | -0.110 | -0.165/-0.057 |
| lAG → mPFC | +0.138 | 0.065/0.211 | +0.105 | 0.031/0.180 |
| lAG → lAG | -0.249 | -0.394/-0.105 | -0.228 | -0.371/-0.085 |
| lAG → lAI | -0.109 | -0.189/-0.029 | -0.115 | -0.196/-0.035 |
| lAG → rAI | -0.177 | -0.289/-0.065 | -0.137 | -0.248/-0.026 |
| lAG → lFEF | -0.123 | -0.216/-0.031 | -0.154 | -0.250/-0.058 |
| rAG → rAG | +0.153 | 0.143/0.293 | +0.202 | 0.061/0.344 |
| dACC → PCC | +0.178 | 0.064/0.292 | +0.160 | 0.042/0.278 |
| dACC → mPFC | +0.161 | 0.088/0.234 | 0.000 | 0.000/0.000 |
| dACC → lAI | +0.123 | 0.032/0.214 | +0.096 | 0.003/0.190 |
| dACC → lIPS | -0.126 | -0.208/-0.045 | -0.176 | -0.258/-0.094 |
| lFEF → PCC | -0.182 | -0.278/-0.085 | -0.111 | -0.206/-0.017 |
| lFEF → mPFC | -0.235 | -0.331/-0.140 | -0.247 | -0.345/-0.150 |
| lFEF → rAG | -0.144 | -0.241/-0.046 | -0.127 | -0.223/-0.031 |
| lFEF → lAI | -0.188 | -0.313/-0.063 | -0.217 | -0.344/-0.090 |
| lFEF → rIPS | +0.168 | 0.063/0.274 | +0.157 | 0.048/0.266 |
| rFEF → lAG | +0.151 | 0.079/0.224 | +0.209 | 0.135/0.283 |
| lIPS → PCC | +0.236 | 0.130/0.342 | +0.164 | 0.056/0.272 |
| lIPS → lAG | +0.107 | 0.026/0.188 | +0.168 | 0.087/0.249 |

| | | | | |
|---|---|---|---|---|
| lIPS → rAG | +0.128 | 0.063/0.193 | +0.095 | 0.033/0.157 |
| rIPS → lAG | -0.116 | -0.179/-0.054 | -0.135 | -0.195/-0.075 |
| rIPS → rIPS | +0.237 | 0.102/0.371 | +0.221 | 0.086/0.357 |

*Table S3. Changes to between regions effective connectivity signifying lasting effects of LSD. Only the change from placebo > 0.1 Hz with effective connectivity change < 0.1Hz in the later effects at 300 min, compared to 75 minutes, are reported. Values reported in Hz. All results are for posterior probability > 0.99.*

#### 4. Hierarchical Connectivity Strength Calculation

Average efferent and afferent connectivity for each network, indicates hierarchical organisation of the 3 networks. It remains to be clarified that the "hierarchy" presented here is a relationship between intrinsic networks, characterized by the differences between efferent and afferent connectivity strength. The higher ranking only means greater efferent–afferent difference, rather than higher ranking in cortical or functional hierarchy - this interpretation is applied to describe hierarchy between networks [9].

Calculations for the three networks across placebo, 75 minutes post administration and 300 minutes post administration:

*Placebo*

SN (0.21 +0.13) - (0.16+0.42) = .27 - .42 = -0.15

DMN (0.69 + 0.16) - (0.35+0.21) = 0.85 - 0.56 = 0.29

DAN (0.42 + 0.35) - (0.69 +0.13) = .77 - .82 = -0.05

*75 minutes post administration*

SN (0.22 +0.13) - (0.73+0.86) = .35 - 1.59 = -1.24

DMN (0.51 + 0.73) - (0.31+0.22) = 1.24 - 0.53 = 0.71

DAN (0.86 + 0.31) - (0.51 +0.20) = 1.17 - .71 = 0.46

*300 minutes post administration*

SN (0.01 +0.57) - (0.71+0.66) = .58 - 1.37 = -0.79

DMN (1.29 + 0.16) - (0.35+0.21) = 0.85 - 0.56 = 0.29

DAN (0.66 + 0.23) - (1.29 +0.57) = .89 - 1.86 = -0.97
